# Economic burden and catastrophic health expenditure associated with COVID-19 hospitalisations in Kerala, South India

**DOI:** 10.1101/2021.12.20.21268081

**Authors:** Ronnie Thomas, Quincy Mariam Jacob, Sharon Raj Eliza, Malathi Mini, Jobinse Jose, A Sobha

**Affiliations:** Government Medical College, Kottayam; Pushpagiri Institute of Medical Sciences and Research Centre, Thiruvalla; Kasturba Medical College, Mangalore

## Abstract

**Introduction:** Catastrophic health expenditure during COVID-19 hospitalization has altered the economic picture of households especially in low resource settings with high rates of COVID-19 infection. This study aimed to estimate the Out of Pocket (OOP) expenditure and the proportion of households that incurred catastrophic health expenditures due to COVID-19 hospitalisation in Kerala, South India.

**Materials and Methods:** A cross-sectional study was conducted among a representative sample of 155 COVID-19 hospitalised patients in Kottayam district over four months, using a pretested interview schedule. The direct medical and non-medical costs incurred by the study participant during hospitalisation and the total monthly household expenditure were obtained from the respective COVID-19 affected households. Catastrophic health expenditure was defined as direct medical expenditure exceeding 40% of effective household income.

**Results:** From the study, median and mean Out of Pocket (OOP) expenditures were obtained as USD 93.57 and USD 502.60 respectively. The study revealed that 49.7% of households had Catastrophic health expenditure, with 32.9% having incurred Distress financing. Multivariate analysis revealed being Below poverty line, hospitalisation in private healthcare facility and presence of co-morbid conditions as significant determinants of Catastrophic health expenditure.

**Conclusion:** High levels of Catastrophic health expenditure and distress financing revealed by the study unveils major unaddressed challenges in the road to Universal health coverage.

## 1. Introduction

The second wave of COVID-19 in India was a humanitarian and public health crisis which has overwhelmed the country’s healthcare system (Asrani et al., 2021). The newly emerged Delta B.1.617 variant with increased transmissibility and pathogenicity was behind the devastating turn the pandemic took in April 2021 in India (Cherian et al., 2021; Thangaraj et al., 2021). As of October 1, India had reported more than 3.37 million confirmed COVID-19 cases and 448,605 deaths (#IndiaFightsCorona 2021.). COVID-19 continues to spread across the southern state of Kerala with significant health consequences and mortality (Ellis-Petersen, 2021; Thomas et al., 2021). In addition to the direct effects of COVID-19, the economic and social disruption associated with the pandemic has had a substantial impact on the livelihood of the population and has exposed the healthcare disparities existing in socially disadvantaged communities (El-Khatib et al., 2020; Liao & De Maio, 2021). COVID-19 patients from such communities disproportionately experienced the financial burden due to hospitalisations (Liao & De Maio, 2021). The immediate apparent reason for this healthcare disparity is the underlying economic inequality.

Inadequate financial risk protection leads to high out-of-pocket expenditure (OOPE) on healthcare. The OOP contribution reaches almost three-quarters of total expenditure on health in India (van Doorslaer et al., 2007). The Twelfth Five Year Plan of India had reduction of Out Of Pocket expenditure (OOP) as one of the measurable targets (Planning Commission of India. 2013). NITI Aayog’s report on the need for Health insurance coverage states that the increased reliance on the private healthcare sector, makes the population vulnerable to increasing out of pocket payments (Sarwal & Kumar, 2021). Evidence on OOPE is important to protect vulnerable poor households from catastrophic health expenditures (CHE) by providing appropriate financial risk protection. Health spending is considered to be catastrophic when a household must reduce its basic expenditure over a period of time to cope with health costs (Xu et al., 2003a). Catastrophic healthcare spending is considered as an indicator of financial protection under Universal Health Coverage for the Sustainable Development Goals (United Nations., 2017.; Wagstaff et al., 2018). In Africa and Asia the number of people incurring catastrophic payments increased in the last decade (Wagstaff et al., 2018). As pointed out by Xu et al (2003a), policy makers need better understanding of any socioeconomic, cultural or demographic characteristics that make people more vulnerable to catastrophic payments. Furthermore the economic weakening of a household as a result of health care expenses can lead to distress financing manifesting as borrowing money, mortgaging gold and selling of assets. Indicators like CHE do not consider the economic impact of health conditions like COVID-19 beyond the health sector (Boccuzzi, 2003). As suggested by Wagstaff et al (2018), health care-seeking has an opportunity cost beyond the direct medical costs in the form of productivity losses. This is especially important in the context of COVID-19 pandemic as the income loss can either be due to temporary reasons such as quarantine or job absenteeism and permanent causes like mortality or disability.

To the best of our knowledge, no study from India has systematically examined the economic burden of COVID-19 patients who required in-patient care. Evidence on the scale of hospitalisation expenditures would enable decision-makers to optimize the allocation of scarce healthcare resources. With this background we conducted a cross sectional study to primarily estimate the Out of Pocket (OOP) expenditure and the proportion of Catastrophic Health Expenditures (CHE) among hospitalised patients with COVID-19. Secondly we assessed the proportion of households facing impoverishment or distress financing due to catastrophic payments.

## 2. Materials & Methods

### Study sample

The study population included patients of all age groups and genders diagnosed with COVID-19 and required in-patient care in Kottayam district from May to August 2021. The sampling frame was the line list of patients diagnosed as COVID-19 using either RT-PCR or Rapid antigen test and admitted in any private hospitals, Covid second line treatment centres (CSLTC) or designated Covid hospitals in Kottayam district. The authors performed a systematic random sampling for selecting study participants from the linelist, which also included patients expired at the time of data collection for whom details were collected from the next of kin. The investigators telephonically collected the data using a pre-tested structured interview schedule. Verbal informed consent was obtained from all study participants. Pregnant patients were excluded and in cases of multiple hospital admissions for a single patient, the last hospital of admission was considered for study. The minimum sample size required was calculated to be 149.

### Estimation of costs

As participants in household surveys are generally reluctant to reveal their total income, we used monthly household final consumption expenditure as the proxy for monthly effective income and the food expenditure as a proxy for household subsistence expenditure as explained by Xu et al (2003b). Expenditure data were collected as reported by the participants and the hospitalization expenditures of participants who were admitted in private hospitals were verified by the copies of available hospital bills sent electronically through Whatsapp or email. The investigators calculated the direct medical costs per COVID-19 patient as the sum of the inpatient cost in each categories namely room/ICU rent, physician consultations, nursing services, drugs, medical consumables, imaging, electrocardiography, laboratory services, invasive and non-invasive ventilation, blood products transfusion and other procedures or interventions like hemodialysis. The direct non medical costs for each COVID-19 patient was calculated as the sum of expenses incurred for food, travel and accommodation for the patients and the bystanders. Out-of-Pocket Expenditure (OOPE) was taken as the sum of direct medical and direct non medical expenses paid by the house-hold at the time of hospitalisation for COVID-19 after deducting for the aid received for financial risk protection. For defining Catastrophic health expenditure we used the threshold and approach reported by Xu et al (2003a). This approach uses the households capacity to pay which is a measure of the non-subsistence income of the household calculated as effective income minus subsistence income. As per the definition we used, a household is said to incur catastrophic health expenditures when out-of-pocket spending on COVID-19 hospitalisation exceeds 40% of the household’s capacity to pay (Xu et al., 2003a). Distress financing was said to occur when the household is forced to borrow money or to take a personal loan or sell/mortgage land,gold or other assets for meeting healthcare expenditure during hospitalisation for covid 19 (Joe, 2015).The lost productivity was quantified only for the earning members of the household. In addition to income loss due to death, disability and job absenteeism, we included income loss due to isolation, quarantine or patient care for any earning member of the household for quantifying the indirect costs. (Jo, 2014). The friction period for estimating loss of income from permanent loss of productivity was fixed at 3 months.

### Data Analysis

The outcome measures were the median OOP Expenditure, proportion of households incurring Catastrophic health expenditure, proportion of households experiencing distress financing and the proportion of households facing temporary and permanent productivity losses. The households were divided into income tertiles based on the effective income. We used the Mann Whitney U test and the Kruskal-Wallis test for nonparametric analyses of OOP expenditures. To evaluate the correlates of catastrophic health spending we created a binary logistic regression model. Variables were introduced into the regression model if they were found to be associated with CHE at p < 0.20. All statistical analyses were conducted using the statistical software SPSS version 20.0 (IBM Corp, Armonk, NY). We used a conversion rate of USD$ 1 = 74.9 INR. The Institutional Review Board (IRB)of Government Medical College, Kottayam approved this study vide reference no 94/21.

## 3. Results

The authors interviewed 179 patients or next of kin, of whom 155 fulfilled the inclusion criteria. The mean age of the study participants was 55.45 ±19.93 years with a female predominance(55.5%). The average household size was 4.21±1.57 persons. Among the study participants, 38.7% of belonged to households with percapita income below the international poverty line and 12.3% were unemployed. The mean duration of hospitalisation was 11.35±10.75 days. Figure 1 shows the summary of assessment results with regard to the objectives of the study.

**Figure 1.**
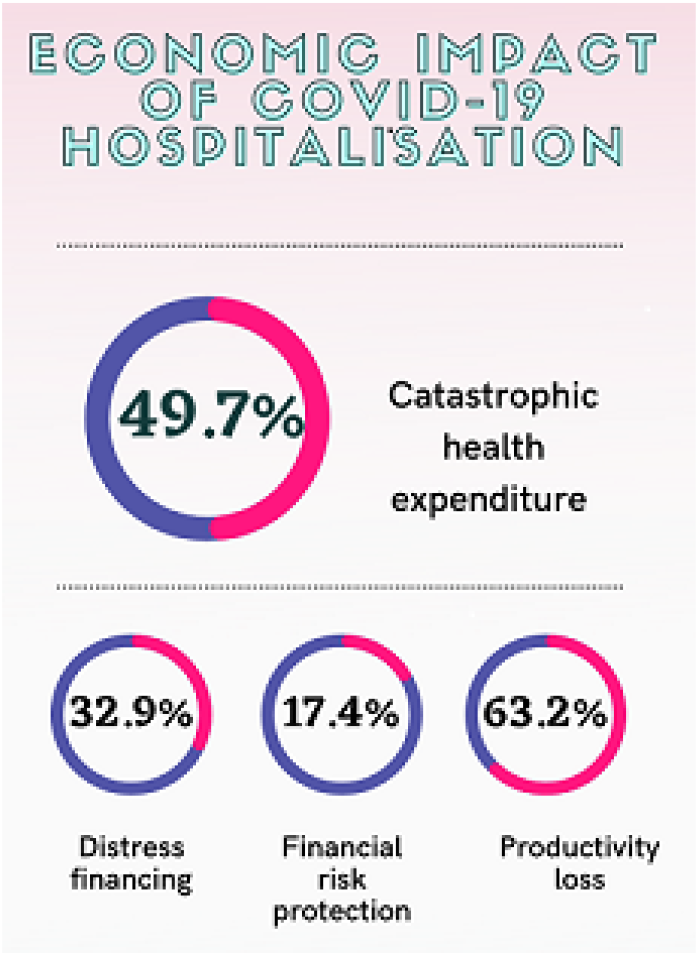
Summary of assessment results.

The present study found that the median (IQR) total out-of-pocket expenditure (OOP) among the study participants was INR 7010 (1500-38150), USD 93.57. The median (IQR) OOP expenditure for inpatient treatment of COVID-19 in public and private hospitals were INR 2980 (825 – 8850),USD 39.78 and INR 38100 (10237.5 - 63625),USD 508.67, respectively.

Our analysis revealed that a total of 49.7 % (95%CI 41.9, 58.1) of households incurred catastrophic health expenditure due to COVID 19 hospitalisation. For the patients who underwent treatment in public sector institutions, 37.6% of households experienced catastrophic health expenditures. Whereas, for private sector hospitalisations the health expenditure was catastrophic for 64.3% households. As Shown in Table 1, the median OOP expenditure was found to be significantly higher in patients with advancing age, comorbidities, severe disease and bad clinical outcomes. Significant variation of OOP expenditure was observed across the different income tertiles for COVID hospitalisations.

**Table 1.** The median (IQR) Out of Pocket Expenditures

| Correlates | OOP Expenditure |  | p value |
| --- | --- | --- | --- |
|  | Median (IQR), INR | Median, USD |  |
| **Age** |  |  | **0.048** |
| <= 45 years | 7490 (1520- 57000) | 100 |  |
| > 45 years | 21035 (3225-50450) | 280.84 |  |
| **Gender** |  |  | **0.267** |
| Male | 9015 (2010-49800) | 120.36 |  |
| Female | 24980 (2950-53000) | 333.51 |  |
| **Income tertile** |  |  | **0.017** |
| Upper | 42200 (18800-60650) | 563.41 |  |
| Middle | 5550 (1350-62750) | 74.09 |  |
| Lower | 6525 (1875-27010) | 87.11 |  |
| **Health Facility** |  |  | **<0.001** |
| Government | 2980 (825 - 8850) | 39.78 |  |
| Private | 38100 (10237.5 - 63625) | 508.67 |  |
| **Comorbidity** |  |  | **0.043** |
| Present | 19325 (3725-53040) | 258.01 |  |
| Absent | 8990 (1025-48550) | 120.02 |  |
| **Severity of Illness** |  |  | **0.001** |
| Mild- Moderate | 7240 (2005-46575) | 96.67 |  |
| Severe | 31000 (15100-111200) | 413.88 |  |
| **Clinical Outcome** |  |  | **0.018** |
| Death | 18200 (7010- 49500) | 242.98 |  |
| Cured | 5000 (1362.5- 31900) | 66.75 |  |
| **Financial Protection** |  |  | **<0.001** |
| Insured | 2400 (250-6490) | 32.04 |  |
| Not insured | 14030 (2225- 45100) | 187.31 |  |

The median out-of-pocket costs for COVID-19 admissions are similar regardless of gender, educational status and occupational categories. In approximately one third of the hospitalizations (32.9%), patient costs were covered through borrowing, gold loans or sale of assets. Only 27 (17.4%) participants reported utilising health insurance plans. The significant findings with regard to the odds of catastrophic health expenditure associated with COVID-19 hospitalisations are summarised in Table 2.

**Table 2.**
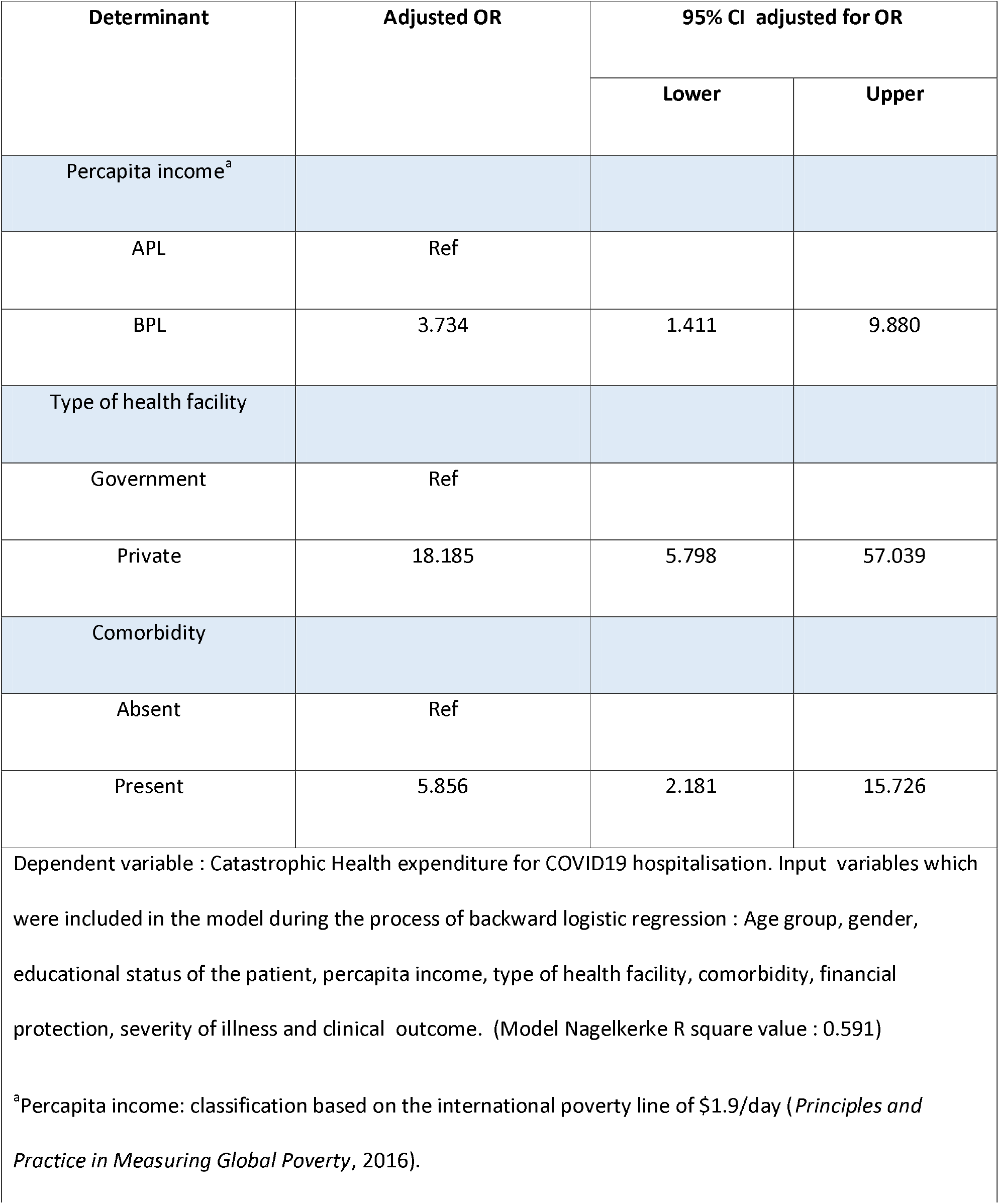
Binary logistic regression results of determinants of Catastrophic Health expenditure on COVID 19 hospitalisation.

| Determinant | Adjusted OR | 95% CI adjusted for OR |  |
| --- | --- | --- | --- |
|  |  | Lower | Upper |
| Percapita income<sup>a</sup> |  |  |  |
| APL | Ref |  |  |
| BPL | 3.734 | 1.411 | 9.880 |
| Type of health facility |  |  |  |
| Government | Ref |  |  |
| Private | 18.185 | 5.798 | 57.039 |
| Comorbidity |  |  |  |
| Absent | Ref |  |  |
| Present | 5.856 | 2.181 | 15.726 |

Dependent variable : Catastrophic Health expenditure for COVID19 hospitalisation. Input variables which were included in the model during the process of backward logistic regression : Age group, gender, educational status of the patient, percapita income, type of health facility, comorbidity, financial protection, severity of illness and clinical outcome. (Model Nagelkerke R square value : 0.591)
<sup>a</sup>Percapita income: classification based on the international poverty line of \$1.9/day (*Principles and Practice in Measuring Global Poverty*, 2016).

A total of 98 (63.2%) households reported productivity loss, of which majority (48.4%) reported temporary income loss. 14.8% of the households reported permanent income loss for atleast one member.. The mean indirect expenditure for the study participants was found to be INR 18830.9 ± 29424.

## 3. Discussion

In the present study we report the healthcare expenditure data on COVID-19 hospitalisations as the median OOP expenditures, the proportion of households facing CHE and the proportion of households where OOP was sourced by distress financing. The overall median (IQR) OOP expenditures on COVID-19 hospitalisations in central Kerala were INR 7010 (1500-38150), USD 93.57. This was the lowest reported OOP expenditure for in-patient care compared to those reported elsewhere. Evidence on costs of COVID-19 hospitalisations to date are drawn from high income countries. According to a report on the COVID-19 hospitalization cost from Spain, the mean cost per patient was USD 12,108 (Carrera-Hueso et al., 2021). Analysis based on co-payments data from the United States found the mean total out-of-pocket spending to be USD 3840 for privately insured patients and USD 1536 for patients with medicare services (Chua et al., 2021). The estimates from China puts the medical cost as USD 6827 per treated case while the mean daily hospitalization cost per patient for COVID-19 care in Saudi Arabia was reported to be USD 1384.57(Li et al., 2020) (Khan et al., 2020).

We observed that the median OOP expenditure was nearly ten times higher (INR 38100, USD 508.67) in private as compared to the public sector (INR 2980, USD 39.78). COVID-19 has a wide spectrum of clinical presentations and the OOP expenditure was found to vary across severity categories. It was also observed that the median OOP payments for severely ill patients and patients who died were significantly higher. This was in agreement with the European Centre for Diseases Prevention and Control country summary data which when adjusted for the GDP income category, found a positive association between OOP expenditure for COVID hospitalizations and mortality (El-Khatib et al., 2020). When comparing the results of the present study to the costs incurred during hospitalisation for other respiratory infections in India, it was observed that the out of pocket costs were substantially higher for Covid care (Kumar et al., 2019). This observation was in contrast to findings from studies reported elsewhere. An analysis based on the administrative claims data among the patients admitted for pneumonia between 2016 and 2019 in the United States estimates the mean out-of-pocket spending as USD1,961 (Eisenberg et al., 2020). A systematic review on the health care costs of influenza like illness in high income countries range from US$19 in Korea to US$323 in Germany (Federici et al., 2018). Though the expenses associated with general illness is not comparable with COVID-19, studies based on the National Sample Survey data report the mean OOP expenses on hospitalization for all causes to be INR 19,210. (Kastor & Mohanty, 2018)

We found that a total of 77 (49.7%) out of the 155 eligible households studied incurred catastrophic health expenditure due to hospitalisation for COVID-19. Wagstaff et al (2018) reported the global incidence of catastrophic expenditure, at a threshold of 10% income, as 11·7% in the year 2010. A multi-country analysis on the magnitude and distribution of OOP payments by van Doorslaer et al (2007) estimates the incidence of catastrophic payments in India, defined at 25% of non-food expenditures, to be nearly 10% in the year 2007. There are considerable variations observed in the CHE estimates from national surveys done across India (Raban et al., 2013). Research work based on the National Sample Survey data done in 2014 demonstrated that 24.9% of the Indian households incurred CHE (Pandey et al., 2018). This survey reports an increase of 17.0% in CHE from that in 2004. The World Health Survey (WHS) done in the year 2003 reported that 33.9% of households in India suffered CHE. These variations are attributed to the differences in the survey methodologies used (Raban et al., 2013). The results of our regression analyses highlight the link between CHE and percapita income. This finding further substantiates the importance of addressing the problem of income inequality which can only be resolved by providing universal health coverage and social security benefits intended to protect households from income loss associated with hospitalisation. But with the government health spending standing at 1·28% of GDP, inadequate public expenditure is going to be the greatest challenge in achieving universal health care(Bang, 2015; MOHFW, 2018.). Our study also demonstrates a robust relationship between the type of health facility where the patient sought care and CHE. A report published in the year 2021 claims that more than half of Kerala’s population rely on the public sector for episodes of acute illness (Nair & Varma 2021). The state government took a policy decision to offer publicly financed decentralized medical care services to overcome the financial barriers to accessing COVID-19. This approach taken by the state government led to the establishment of district level first line and second line covid care centres care which provided free treatment and accommodated a major share of hospitalizations. Our finding is consistent with the observations in other studies that have found a similar health seeking pattern. The 75^th^ round of National Sample Survey (NSS) carried out in the year 2017 reports the average medical expenditure incurred during Hospitalisation was more than seven fold in a private hospital than in a government hospital (MOSPI, 2019). The analysis from the present study adds to a group of studies that have established that comorbid conditions are significantly associated with increasing medical costs for hospitalisations for COVID-19 (Carrera-Hueso et al., 2021; Chua et al., 2021; Miethke-Morais et al., 2021). Even though the existing body of evidence from low and middle income countries emphasizes on the role of health insurance in protecting households from catastrophic health costs,the present study did not identify such an association in the regression model (Huffman et al., 2011). Recent research has demonstrated that coverage under the public health insurance programs for those below poverty line had no significant influence on decreasing OOP payments. Majority of the participants with financial protection in the study sample were privately insured. We observed a low utilisation rate for Ayushman Bharath, the largest publicly financed health insurance scheme, with only 4 participants reporting availing the service.This could be attributed to the slow enrolment rate and relatively lower number of empanelled private hospitals for the scheme in the state.

The estimation of the proportion of distress financing in the study population is imperative in understanding the sources of OOP payments for COVID-19. The present study demonstrates that one-third of the study participants(32.9%) resorted to borrowing of money, private gold loans and selling of assets for meeting the hospitalisation expenses. This is in line with peer reviewed evidence from India. Joe et al found that distress financing account for 58 and 42% share of the total OOP payments for hospitalised care in rural and urban India, respectively (Joe, 2015). Among the people suffering from the burden of cancer in India, distress financing occurred in 43% of the population. (Kastor & Mohanty, 2018).

This is consistent even in the case of maternal services in India. In spite of the cash incentives for institutional delivery since the implementation of the National Health Mission, a recent study found that one in four mothers in India resorted to borrowing or selling to meet the OOP expenses on institutional delivery. (Mishra & Mohanty, 2019).

Our study has several limitations. Firstly it was not possible for us to correctly analyse the cost components in the OOP expenses as patients who sought care in public health facilities were not provided a hospital bill or expenditure statement. Secondly recall bias may have affected the retrospective assessment of the expenditures. Thirdly the approach of calculating the monthly household income based on total expenditure may have underestimated the actual household income. Notwithstanding these limitations, the study provides key information that will enable policy makers to prioritise interventions to ensure health equity and universal health coverage.

## 4. Conclusion

The present study provides an insight into the extent to which COVID-19 hospitalisations have disrupted household economies. Though the study population had one of the lowest reported OOP payments for COVID-19 hospitalisations worldwide, nearly half of them suffered catastrophic expenditures which led to impoverishment in approximately a third. From the study findings, it is plausible that catastrophic payments would prevent patients under home isolation for COVID-19 from seeking necessary health care. Given the current situation, the government should take efforts to scale up coverage and implementation of social health insurance programmes to extend the social safety net. The findings from the study reiterated the need for such major reforms in health financing systems. Further research on the wider economic consequences of the COVID-19 pandemic would be essential for the attainment of universal health coverage in the context of unforeseen health crises across the world in the years to come.

## Data Availability

All data produced in the present work are contained in the manuscript

